# Health economic model assumptions of pharmaceutical treatment paths compared with real-world evidence for patients with type 2 diabetes: A nationwide cohort study

**DOI:** 10.1101/2025.06.20.25329913

**Authors:** Henrik Vitus Bering Laursen, Flemming Witt Udsen, Morten Hasselstrøm Jensen, Peter Vestergaard, Søren Paaske Johnsen

## Abstract

2

**OBJECTIVE:** The glucagon-like peptide-1 (GLP1) and sodium-glucose co-transporter-2 (SGLT2) classes are increasingly being used around the globe to treat type 2 diabetes and its comorbidities. Decision-analytical models (DAMs) are used to evaluate the cost-effectiveness of the different products within these classes, but their assumptions regarding the treatment pathways and time until insulin may not reflect real-world practice. This study compares the model assumptions found in a recent review with registry data.

**RESEARCH DESIGN AND METHODS:** Real-world practice was represented by a nationwide registry-based cohort of 62,238 people with type 2 diabetes included in an early (2012 to 2018), and a late period (2019 to 2021). Treatment pathway assumptions were compared using counts and proportions. Time until insulin initiation was compared among groups starting on the comparators dipeptidyl peptidase-4 inhibitor (DPP4), GLP1, or SGLT2. Incidence rates, hazard ratios, and absolute risks were utilized. The latter were derived from cumulative incidence functions calculated using the Aalen-Johansen estimator, accounting for competing risks.

**RESULTS:** The treatment pathway assumptions of initiating insulin after a short time on the comparators did not correspond with our data. Neither did the assumptions surrounding time until insulin, as the time differed between comparator group, and inclusion period, with SGLT2 having the lowest risk. Further, time until insulin was observed to be much longer in real-world practice than in the model assumptions.

**CONCLUSIONS:** Key model assumptions used to inform decision-makers on the cost-effectiveness of expensive drug classes like GLP1 and SGLT2, are flawed and need to be updated.

---

Type 2 diabetes progression notably increases hospitalizations and mortality, diminishes quality of life, and reduces productivity, partly due to complications associated with the disease (1,2). Since 2006, the low-cost and highly effective metformin has been the standard first-line non-insulin anti-diabetic drug (NIAD) (3). The use of sodium–glucose cotransporter 2 (SGLT2) inhibitors and glucagon-like peptide 1 (GLP1) receptor agonists have expanded greatly in the last 5-10 years (4), and SGLT2 and GLP1 have been strongly recommended since 2018 (5). Recent guidelines (6) started recommending them as first-line options for patients with type 2 diabetes who exhibit cardiorenal risk factors. However, in most cases, the cost of SGLT2 and GLP1 are significantly more expensive than metformin (7,8). Recent research has indicated that significant price reductions are required to establish these treatments as cost-effective first-line options (9).

A standard method for analyzing trade-offs between costs and health outcomes across treatment alternatives is through cost-effectiveness analyses (CEAs) using decision-analytical modelling (DAM). The output of these DAMs influence decision-making regarding use and reimbursement. (10,11) Although results DAMs must be interpreted within their contexts, the consensus of several reviews of CEAs comparing diabetic pharmaceuticals is that SGLT2 and GLP1 are cost-effective compared to sulfonylurea, thiazolidinediones, dipeptidyl peptidase 4 inhibitors (DPP4), insulin, and alpha-glucosidase inhibitors for second-line treatment. (12–18). A recent review conducted by Laursen et al. in 2023 (18) focused on CEAs that have used DAM to compare the DPP4, GLP1, and SGLT2 classes, and found that determining cost-effectiveness heavily relying on two core study assumptions common across many of the included studies. First, a common treatment pathway for patients, called the standard pathway in this paper, was usually assumed. In this pathway, patients were initially prescribed metformin, followed by the addition of the drugs compared in the DAM (DPP4, GLP1, or SGLT2) and ending in the prescription of insulin, either as an addition or a replacement of the comparator. Second, time until insulin initiation varied between three and 13 years across the 23 out of 50 included studies that explicitly reported it, but was most commonly assumed to be three years. (18)

The objective of this study is to evaluate if these two central DAM assumptions are reflected in real-world practice by investigating individuals with type 2 diabetes prescribed with DPP4, GLP1, and SGLT2 as second-line treatments, in terms of their treatment pathways and time to insulin, using real-world data from Danish registries. This investigation could inform decision-makers on how representative or applicable the results of the CEA using DAMs are in a Danish context, as well as inform future models.

## 3 Research Design and Methods

### 3.1 Study Design

To investigate the difference between assumption one (treatment pathways) and assumption two (time to insulin) and real-world data, a register-based cohort of all Danish individuals with type 2 diabetes, who were treated with second-line drugs or beyond, was established. This cohort was divided into a main and a nested cohort. For the main cohort, an overview of common treatment pathways was created by following included individuals from their first prescription of metformin until their last prescription. Comparisons were made between an early (2012-2018) and a late (2019-2021) inclusion period. In the nested cohort, individuals were followed from first exposure to DPP4, GLP1, and SGLT2, and until the first prescription of insulin occurred, death, or end of follow-up.

### 3.2 Setting and data sources

The Danish healthcare system is tax financed with universal healthcare and a long history of creating and maintaining health registries of high quality and validity (20). Danish health registries are linked through a personal identification number, which is given at birth, or upon immigration if one is a resident. Cohort inclusion and the outcomes are based on prescriptions, which were sourced from the Danish National Prescription Database (DNPD) (21). Here, nationwide information on all redeemed prescriptions dispensed by Danish community pharmacies from 1996 can be accessed. Redeemed prescriptions served as inclusion times and as a proxy treatment with the respective drug classes. Data on age and sex was retrieved from the Danish Civil Registration System (19). Finally, hospitalization diagnoses were collected from the Danish National Patient Registry (DNPR), which contains complete nationwide data on hospitalizations since 1978 (19). Data access can be granted if affiliated with a Danish research organization, in the present case through Statistics Denmark.

### 3.3 Study population

The study population included patients with type 2 diabetes between 2012 and 2021. 2012 was the year where all exposure drug classes, DPP4, GLP1 and SGLT2, were available in Denmark, and 2021 marked the end of data availability. To ensure that the patients had reached at least second-line treatment in the inclusion period, they were included if they had received at least two different prescriptions, and at least two prescriptions of NIADs that were different from, and prescribed after, the first. To exclude individuals with possible type 1 diabetes mellitus, any redeemed prescription of insulin (ATC: A10A) prior to, or on, the date of their first NIAD led to exclusion. Additionally, any prescription of NIADs before 2012, and metformin not being among their first drugs, led to exclusion. This cohort was used for finding the most frequently used treatment regimen and comparison of pathways.

For investigating the treatment pathways, a main cohort was constructed whose follow-up started from the first prescription of metformin (ATC: A10BA02) until their last prescription of any NIAD or insulin. Those below 18 years of age were excluded. For investigating time to insulin, a nested cohort was established from the main, with the additional criteria that individuals were followed from the date of receiving the first prescription of their group-defining nNIADs: DPP4 (ATC: A10BH), GLP1 (ATC: A10BJ), or SGLT2 (ATC: A10BK). Follow-up for these individuals ended at first insulin prescription, death, or censoring events which were end of treatment regimen or study period (31st of December 2021). Combination drugs were excluded except for those with DPP4 and metformin (special cases of ATC: A10BD.X), which counted as DPP4. To avoid confusion about nNIAD class group-belonging, those with prescriptions for several drugs among the exposure groups (i.e. both SGLT2 and GLP1), were excluded. Any prescription of insulin before the baseline date also led to exclusion.

### 3.4 Variables

Baseline cohort characteristics included age, sex, inclusion period, diabetes duration, history of CVD, and insulin initiation. To account for changing treatment practices, the cohort was divided into those included before 2019, following the mid-2018 Danish guideline update, assuming a six-month adoption period. Diabetes duration was defined as the time from first prescription of metformin to baseline.

Drug treatment regimens were determined by identifying the drug classes each person was exposed to, ordering them, and establishing criteria for prescription duration. Prescriptions were aggregated by class, so different products within the same class counted as the same regimen. Overlapping prescriptions resulted in multi-drug regimens. Drug classes included sulfonylureas (ATC: A10BB), thiazolidinediones (A10BG), alpha-glucosidase inhibitors (A10BF), DPP4, SGLT2, GLP1, and antidiabetic combination drugs (A10BD).

A drug class was part of a regimen from the first prescription to the last. If the gap between prescriptions exceeded one year, exposure of that drug was paused and resumed with a later prescription. Thus, regimen without drugs in the follow-up period were also possible. The mean time and standard deviation between prescriptions were used to extend the last regimen’s exposure time, assuming treatment continued beyond the last prescription. A single prescription regimen had exposure time equal to the mean plus one standard deviation. Gaps between regimens were closed when the time between them was less than the mean plus one standard deviation. The last treatment date was December 31, 2021.

#### 3.4.1 The standard pathway

*The standard pathway* refers to the commonalities between the patient pathways in the DAMs included in the review by Laursen et al, see below for the conceptual model.

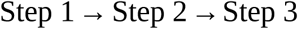

This model outlines the three sequential treatment regimens patients undergo in the DAMs reviewed. The first step is the initial medication, usually metformin or another first-line treatment. The second step involves adding comparators, which marks the start of the simulation. The third step is the final phase, where insulin either replaces or is added to the comparator, typically between three to thirteen years later (18).

To assess external validity, we used the treatment regimen described in *Section 1.4* for two measures: first, the count and proportion of all possible treatment pathways, highlighting the most common; second, the proportion of cohort pathways matching variations of *the standard pathway*. Treatment pathways were arranged in order of occurrence (e.g., “metformin → metformin + GLP1 → insulin”). The proportion of each variation was then presented relative to all pathways experienced by the cohort. The modeled variations are shown below.

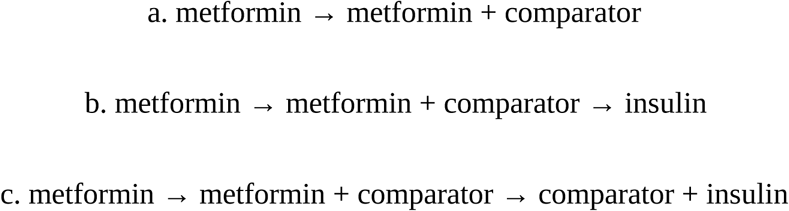

Where *comparator* refers to either DPP4 or GLP1 or SGLT2. In the third step of model *c*, the comparator must be the same as in the second step. These pathways were coded as strict or open. Strict meant that a pathway would only be counted if the exact order of drugs were present. Open pathways were also constructed, where the presence of the pathway was counted as soon as the steps were present. This resulted in a total of 18 distinct pathways.

#### 3.4.2 Time to insulin initiation

The exposures were treatment with the nNIADs DPP4, GLP1, and SGLT2. Insulin initiation was the outcome of the analysis and was defined as any occurrence of a prescription of any insulin class (ATC: A10A.X) within the follow-up period, after the exposure began. Age, diabetes duration, sex, and history of CVD were considered as potential confounders, while the period of inclusion was considered a potential effect modifier. History of CVD was included to control for bias by indication due to the cardio-protective effects of SGLT2 and GLP1. It was defined as any hospitalization occurring prior to baseline, with CVD-related conditions. The diagnostic codes associated with the hospitalizations are detailed in Appendix A1.

### 3.5 Statistical methods

Baseline characteristics are presented for the nested cohort, with means and standard deviations for continuous variables, and counts and proportions for categorical variables. The presence of the pathways in the registry data was evaluated using counts and proportions, for each variation of the pathway, as described in *Section 1.4.1*. The incidence rates (IR) were computed as the number of events per 1000 person years, taking the accumulated person-time at risk into account. Reliability and precision of the IR was assessed by estimating 95% confidence intervals derived from the standard error of a Poisson distribution.

Survival analysis was used to estimate time until insulin initiation, and the analysis was stratified by inclusion period to account for difference in time to insulin between the two inclusion periods. To estimate the difference in time to insulin initiation between the exposure groups, the Aalen-Johansen estimator was used to account for the competing risk of death. Plots were generated to compare the cumulative incidence function (CIF) between the different exposure groups, which provided an estimate of the absolute risk of insulin initiation. The absolute risk of insulin initiation for each exposure group were extracted from the CIF. When there was insufficient follow-up, the equations below were used to predict the risk beyond the follow-up time of the study, until 13 years, which was the upper range of years until insulin was initiated in the included studies in the review by Laursen et al (18).

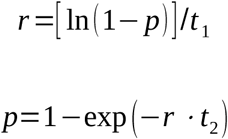

Where *r* is the rate at time *t_1_*, and *p* is the risk of insulin initiation at time *t_2_*. Predictions were based on the last absolute risk estimate, for each exposure group and inclusion period, where at least 10% were still at risk.

The relative hazards of insulin initiation associated with each exposure group were estimated with hazard ratios (HR) derived from the Cox proportional hazards model. DPP4 was chosen as reference. An unadjusted model using only the exposure groups was fitted for each exposure group to estimate crude HRs. The adjusted model included the following covariates: age, diabetes duration, sex, and history of cardiovascular disease (CVD).

Statistical comparison of CIFs between the exposure groups was conducted with Gray’s test. Testing the proportional hazards assumption was conducted by assessing the parallelism of the curves for the different exposure groups on a log-log plot. The assumption was satisfied with approximate parallel curves. The alpha-level for significance was set at 0.05. All statistical analyses, graphs, and summaries were performed using the statistical software *R* (version 4.3.3) and the *survival*, *cmprsk*, and *prodlim* packages.

## 4 Results

### 4.1 Study population

A total of 212,752 individuals receiving NIADs since 2012 were identified, with 62,238 eligible for treatment pathway analysis and 55,430 for time to insulin analysis. The selection process for the two cohorts can be seen in Figure 1 and the baseline characteristics of both cohorts can be found in Appendices A2 and A4.

**Figure 1:**
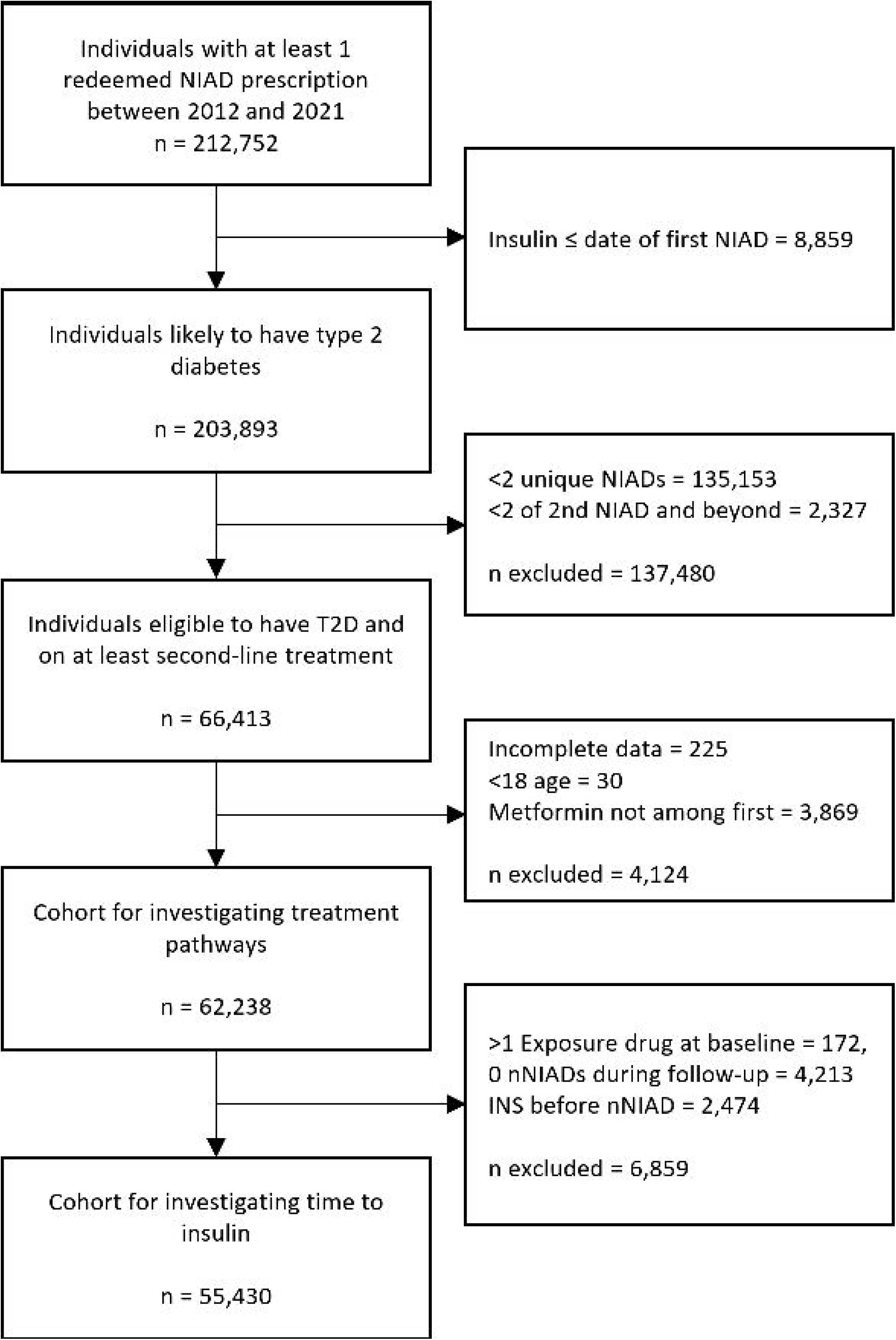
Flow diagram showing the selection of the cohort. NIAD = non-insulin anti-diabetic drug, nNIADs = newer NIADs.

A larger proportion of those aged 18-29 starting on GLP1 were women (Early inclusion [EI]: 7.2% vs 3.6%, late inclusion [LI]: 6.1% vs 1.6%). Men had twice the proportion of cardiovascular disease (CVD) compared to women (EI: 9% to 9.3% vs 4.2% to 6.5%, LI: 7.5% to 13.1% vs 3.4% to 7.5%). The proportion of patients with CVD starting on SGLT2 increased between the two periods (Men: 9% to 13.1%, women: 5% to 7.5%), while it decreased for those starting on GLP1 (Men: 9% to 7%, women: 4.2% to 3.4). SGLT2 users had the lowest proportion of insulin initiation in both periods (EI: 12.4% to 13.3%, LI: 3.1% to 4%), with GLP1 reaching similar levels in the late period (EI: 22.2% to 24%, LI: 2.6% to 3.9%).

### 4.2 The standard pathway

The most common regimen experienced pathway for those included early was “metformin → SGLT2+metformin → metformin” and ocurred for 3.9%. In the late period, it shifted to “metformin → GLP1+metformin → GLP1” (9.5%). From the early to the late time period, presence in a regimen among the top 20 pathways grew most for SGLT2 (12.2% to 30.2%), followed by GLP1 (8.6% to 24.4%). DPP4 presence decreased (6.1% to 4.9%) and SU dissapeared entirely. Although in low proportions, SGLT2 and GLP1 initiation as first-line appeared in the top 20 pathways in the late period (SGLT2: 0.79%, GLP1: 0.64%). See Appendix A3 for more details. In *Table 1*, the presence of different variations of *the standard pathway* in the cohort is presented. There was low presence of all the variations of the open pathways, and the closed pathways were almost not represented.

**Table 1:**
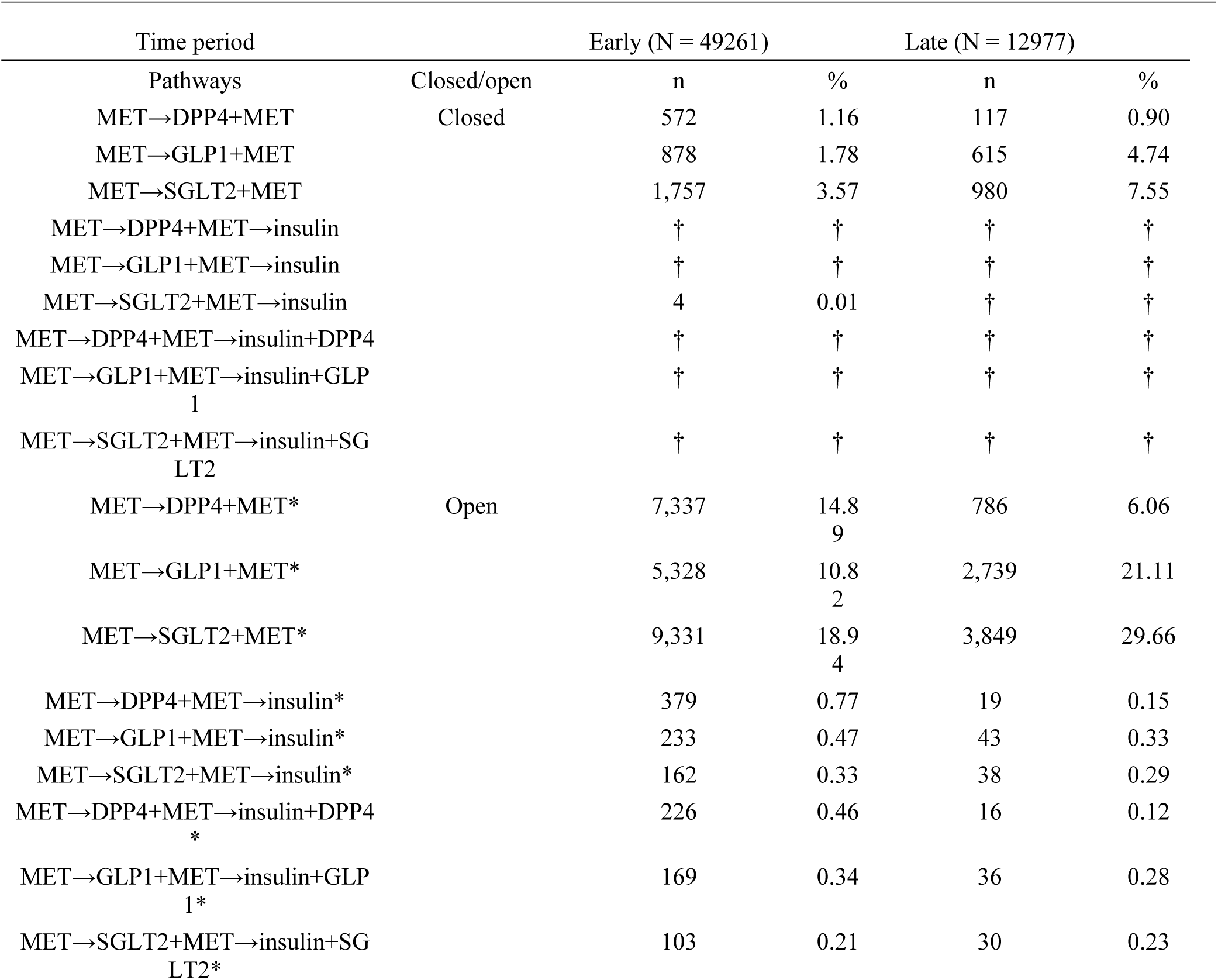
The different variations of the standard pathway, and the counts (n) and proportion (%) of how many in the cohort experienced them out of all pathways, as well as the total amount (N) included in the cohort in each period. * = The pathway is counted if it has any combination of pathways as long as it follows what precedes the “*”, † = Numbers too low to safely export, or 0. MET = Metformin, DPP4 = dipeptidyl peptidase-4 inhibitor, SGLT2 = sodium-glucose cotransporter 2 inhibitor, GLP1 = glucagon-like peptide 1 receptor agonist.

### 4.3 Time to insulin

#### 4.3.1 Aalen-Johansen estimates

In Figure 2, the curves for the CIF are shown. In the early inclusion period, the absolute risk of insulin initiation at 3 years was 15.1% for DPP4, 16.4% for GLP1, and 9.8% for SGLT2. By the late inclusion period, these risks decreased to 13% for DPP4, 8.9% for GLP1, and 7.8% for SGLT2. At 13 years, the predicted absolute risk for the early period was 46.6% (DPP4), 51.1% (GLP1), and 35% (SGLT2), while in the late period, the risks dropped to 45.3% (DPP4), 33.3% (GLP1), and 29.8% (SGLT2). Gray’s test confirmed statistically significant differences between drug classes. Appendix A5 contains predicted times until 13 years.

**Figure 2:**
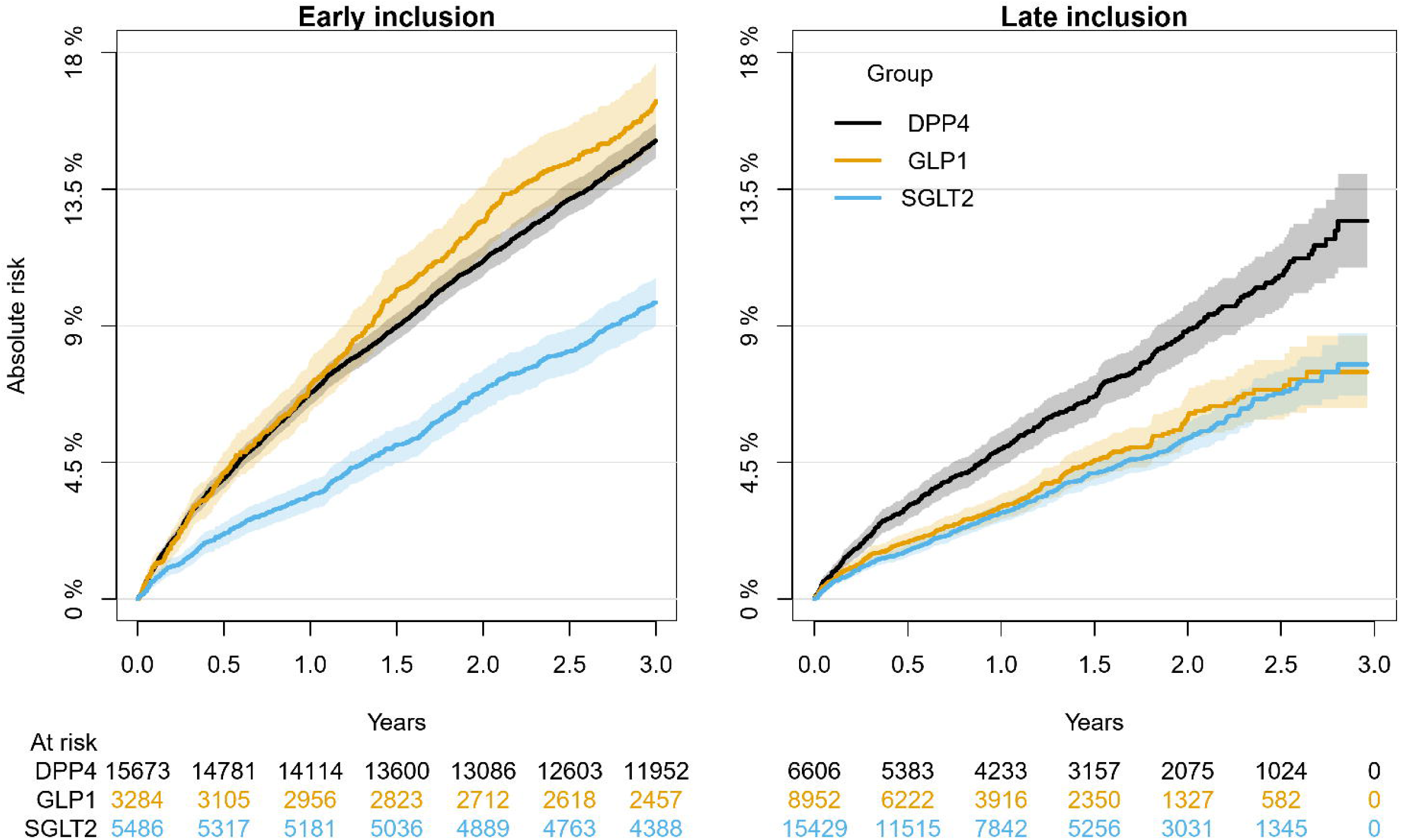
The plot for the Aalen-Johansen cumulative incidence function with 95% confidence intervals. Stratified by inclusion period. Truncated at 3 years for better comparability.

#### 4.3.2 Incidence rates and hazard ratios

The crude IR, unadjusted and adjusted HR for each of the exposure groups within each inclusion period is reported in *Table 2*, while the full model can be found in Appendix A6. In the early inclusion period, the GLP1 group had the highest incidence rate (IR: 56.4, 95% CI [52.4-60.4]), followed by DPP4 (IR: 50.3, 95% CI [48.6-52]) and SGLT2 (IR: 34, 95% CI [31.5-36.5]). By the late period, DPP4 had the highest rate (IR: 48.1, 95% CI [43.7-52.5]), followed by GLP1 (IR: 31.7, 95% CI [28.1-35.4]) and SGLT2 (IR: 28.8, 95% CI [26.3-31.3]). In the early period, starting on GLP1 was associated with a higher risk of insulin initiation (HR: 1.1, 95% CI [1-1.2]) compared to SGLT2 (HR: 0.7, 95% CI [0.6- 0.7]), with DPP4 as the reference. This difference decreased in the late period, with both GLP1 (HR: 0.7, 95% CI [0.6-0.9]) and SGLT2 (HR: 0.6, 95% CI [0.5-0.7]) showing lower risks.

**Table 2:**
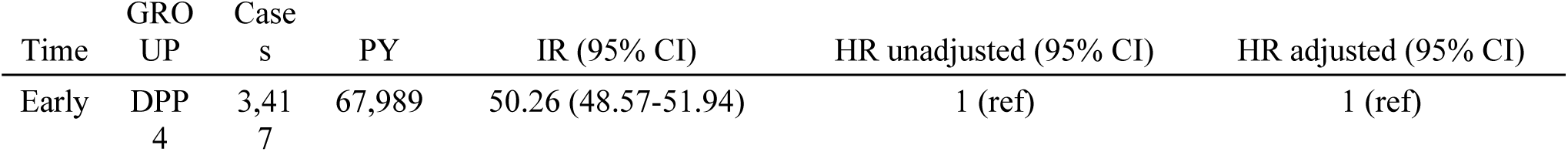
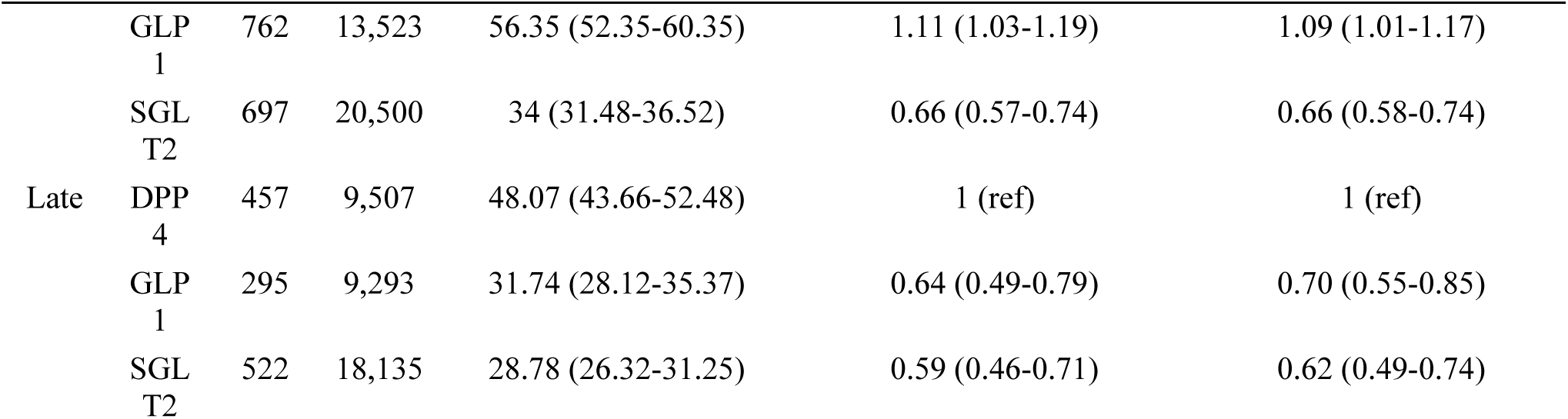
Cases of insulin initation in the cohort, stratified by inclusion period, as well as the effect estimates of the exposure groups, adjusted for sex, age, history of cardiovascular disease, and diabetes duration. PY = Person years, IR = Incidence rate, CI = Confidence interval, HR = Hazard ratio, DPP4 = Dipeptidyl peptidase-4 inhibitors, GLP1 = Glucagon-like peptide-1 receptor agonists, SGLT2 = Sodium glucose co-transporter-2, Late = binary indicator for inclusion before (early) or after and including (late) 2019.

## 5 Conclusions

### 5.1 Main findings

Our analysis shows that the assumption of a linear treatment pathway, as represented by *the standard pathway*, does not align with real-world data from a nationwide Danish cohort. While SGLT2, GLP1, and DPP4 are often used as second-line treatments alongside metformin, the direct progression to insulin replacement or addition for all patients within a short timeframe is not reflected in practice.

Instead, real-world pathways are more complex, involving multiple combinations and pauses before insulin is introduced. This indicates that the CEA studies forming the basis of *the standard pathway* have oversimplified the treatment pathways of patients to a degree that severely limits the external validity of their results. Significant changes in treatment practices, especially after the 2018 paradigm shift, further complicate projections. The changes demonstrated between the early and late inclusion periods indicate that using a fixed assumption about treatment pathways risks producing inaccurate representations of treatment pathways and thus costs and effects included in the DAMs, particularly over longer time horizons. If the 10-year time horizon used in our study can show substantial shifts, projecting for 40 to 50 years, as is typical in DAMs, introduces significantly more uncertainty. The results also call into question the assumptions surrounding the timing of insulin initiation and the lack of differentiation in time until insulin initiation based on nNIAD class. The risk of insulin initiation ranged from 7.8% to 15.1% at three years and 29.8% to 51.1% at 13 years. The shape of the CIF curves also suggest a linear increase in risk over time. This challenges the external validity of the time-to-insulin assumption from *the standard pathway*. Across the inclusion periods, there was a change in risk of insulin initiation for the comparison groups. These findings suggest that neither assuming a constant risk nor a similar risk for the comparison groups over long time horizons in the DAMs is inappropriate, given the large and rapid change observed in this study. In our view, the small differences in baseline characteristics of the between the two inclusion periods could not explain it. Additional follow-up is needed to confirm our findings. Howver, it is unlikely that the current DAMs’ assumptions about insulin initiation reflect actual real-world practice, in a Danish context.

### 5.2 Comparison with other studies

There is a significant knowledge gap regarding the transition between the second and third steps of the standard pathway. Our study shows that various drugs are used before insulin is initiated, though we did not explore the duration or dosage. Literature indicates that persistence on diabetes medications varies by class, with metformin having the highest persistence and GLP1 and SGLT2 showing lower rates, with only about half of individuals continuing their prescriptions after one year (22–24). Given this low persistence, it is not surprising that our study shows low proportions of pathways where insulin is initiated immediately after nNIADs.

One study (25) using data from 2010 to 2017 observed that as time progresses, the time to insulin initiation tends to increase. The median time to insulin initiation increased from 1717 days in 2011 to 1917 days in 2017, with patients on SGLT2 taking longer to initiate insulin compared to those on DPP4. The increasing time to insulin may be linked to the recent rise in SGLT2 and GLP1 use (25). The higher risk of insulin initiation for GLP1 in the early period indicated by our study is also seen in another real-world study(26).

### 5.3 Implications and Future Perspectives

Our findings have important implications for CEAs using DAMs to compare nNIADs for treating type 2 diabetes. First, the treatment pathway from nNIAD initiation to insulin is significantly more complex and involves more steps, than what is modeled in *the standard pathway*. As a result, the models’ ability to accurately represent the costs and effects of drugs used in these intermediate steps is low. Second, the time to insulin in *the standard pathway* is underestimated compared to real-world practice, especially considering changes after the 2018 ADA/EASD guidelines. This leads to further misrepresentation of costs and effects, particularly given the price differences between nNIADs. While the assumption of insulin replacement after three years simplifies modeling, it fails to reflect real-world practice and the growing evidence for GLP1 and SGLT2 benefits. Extending this period to 13 years improves alignment with real-world practice but introduces uncertainty due to low drug persistence.

The effect of these discrepancies on the DAMs depends on the unaccounted drugs, combinations, and durations. If only 50% of patients refill SGLT2 or GLP1 prescriptions after one year, as suggested by Nargesi et al. (22), the economic models fail to capture years of beneficial effects, adverse events, and acquisition costs. Although updating models to reflect real-world practice would add to their already considerable complexity and methodological challenges (18,27,28), it is necessary to improve accuracy or, at a minimum, include this uncertainty in sensitivity analyses. Our findings underscore the need for models to better represent current practice, especially with country-specific data.

### 5.4 Strengths and Limitations

Our study benefits from comprehensive, nationwide prescription data, offering a highly accurate reflection of real-world practice and minimizing both selection and information bias. The inclusion criteria were designed to encompass all individuals with type 2 diabetes fitting the profiles reviewed by Laursen et al. (18), while avoiding misclassification bias, such as including women with polycystic ovarian syndrome. Survival analysis addressed time-related bias.

However, there are limitations. Better matching of patient characteristics and consistent follow-up times between the inclusion period strata could have improved comparability between groups. The precision of the selection criteria for the cohort may have decreased over time, due to the changing indications for the nNIADs. An overview of how often insulin was initiated as a replacement or addition could have provided more information on the degree to which use of nNIADs postpone insulin initiation. Longer follow-up would improve predictions of insulin initiation risk.

### 5.5 Concluding remarks

The discrepancies between the assumptions underlying previous CEAs comparing nNIADs for the treatment of type 2 diabetes using DAMs and real-world practice are substantial enough to have misinformed decision-makers regarding the cost-effectiveness of the nNIADs. Real-world data can be used to update these assumptions. However, this is unlikely to reduce the already considerable complexity of these DAMs.

## Supporting information

RECORD statement

## Data Availability

Underlying micro-data are not available, as they are protected on a research server. All relevant aggregate data is made available in the manuscript or appendices.

## 5.6 Acknowledgements

**5.6.1 Funding and assistance**

This study was supported by Steno Diabetes Center North Denmark.

**5.6.2 Conflict of interest**

PV is the head of research at Steno Diabetes Center North Denmark, funded by the Novo Nordisk Foundation. HVBL has received partial funding for his PhD from Steno Diabetes Center North Denmark, which in turn is funded by the Novo Nordisk Foundation, and additional funding from Boehringer Ingelheim. SPJ has received an institutional research grant from Novo Nordisk. MHJ is full-time employee at Novo Nordisk A/S and owns stocks in Novo Nordisk A/S. FWU has no conflict of interest.

**5.6.3 Author contributions and Guarantor Statement**

HVBL was responsible for the conception of the study question, analysis, presenting, and describing data, and writing the manuscript drafts. SPJ, PV, and FWU provided ideas for the conception of the study question, generated ideas regarding focus areas of the analysis, interpreted the results and provided feedback to the manuscript. MHJ has interpreted the results and provided feedback to the manuscript. All authors agree to be responsible for all aspects of the work and have read and approved the final manuscript.

**5.6.4 Prior presentation**

This study has not been presented previously.

## Twitter summary

- The summary should highlight the key finding, insight, or significance of the paper and should be listed on the title page of the manuscript, below the corresponding author’s contact information.

Study questions cost-effectiveness models for GLP1 & SGLT2 comparison. Nationwide real-world data (2012-2021) shows treatment pathways differ from model assumptions, especially on insulin use. Models need updates to better reflect real-world practices and improve decision-making. #Diabetes #HealthEconomics

## Article highlights

- Why did we undertake this study?
- To test the assumptions used in cost-effectiveness analyses comparing newer drugs for type 2 diabetes against real- world practice.
- What is the specific question(s) we wanted to answer?
- Is the model representation of treatment pathways, and the time until they get exposed to insulin, similar to what can be observed in real-world practice.
- What did we find?
- There are large gaps between the health economic model assumptions and real-world practice.
- What are the implications of our findings?
- When possible, models must be updated with contemporary and country-specific estimates regarding treatment pathways and time until insulin, to properly inform decision makers on the value for money of these newer drugs for the treatment of type 2 diabetes.

## References

1. Bommer C, Sagalova V, Heesemann E, Manne-Goehler J, Atun R, Bärnighausen T, et al. Global Economic Burden of Diabetes in Adults: Projections From 2015 to 2030. Dia Care [Internet]. 2018 May [cited 2019 Nov 8];41(5):963–70. Available from: http://care.diabetesjournals.org/lookup/doi/10.2337/dc17-1962

2. Ng CS, Lee JYC, Toh MP, Ko Y. Cost-of-illness studies of diabetes mellitus: A systematic review. Diabetes Res Clin Pract. 2014 Aug;105(2):151–63.

3. Nathan DM, Buse JB, Davidson MB, Heine RJ, Holman RR, Sherwin R, et al. Management of Hyperglycemia in Type 2 Diabetes: A Consensus Algorithm for the Initiation and Adjustment of Therapy: A consensus statement from the American Diabetes Association and the European Association for the Study of Diabetes. Diabetes Care [Internet]. 2006 Aug 1 [cited 2020 Nov 30];29(8):1963–72. Available from: https://care.diabetesjournals.org/content/29/8/1963

4. Pottegard A, Andersen JH, Sondergaard J, Thomsen RW, Vilsboll T. Changes in the use of glucose-lowering drugs: A Danish nationwide study. DIABETES OBESITY & METABOLISM. 2023 APR 2023;25(4):1002–10.

5. Davies MJ, D’Alessio DA, Fradkin J, Kernan WN, Mathieu C, Mingrone G, et al. Management of hyperglycaemia in type 2 diabetes, 2018. A consensus report by the American Diabetes Association (ADA) and the European Association for the Study of Diabetes (EASD). Diabetologia [Internet]. 2018 Oct 5 [cited 2019 Nov 8];61(12):2461–98. Available from: http://link.springer.com/10.1007/s00125-018-4729-5

6. Committee ADAPP. 9. Pharmacologic Approaches to Glycemic Treatment: Standards of Medical Care in Diabetes—2022. Diabetes Care [Internet]. 2022 [cited 2022 May 6];40:S64–74. Available from: https://diabetesjournals.org/care/article/40/Supplement_1/S64/36805/8-Pharmacologic-Approaches-to-Glycemic-Treatment

7. McEwen LN, Casagrande SS, Kuo S, Herman WH. Why Are Diabetes Medications So Expensive and What Can Be Done to Control Their Cost? Curr Diab Rep. 2017 Sep;17(9):71.

8. American Diabetes Association Professional Practice Committee. 9. Pharmacologic approaches to glycemic treatment: Standards of care in Diabetes—2024. Diabetes Care [Internet]. 2023 Dec;47:S158–78. Available from: 10.2337/dc24-S009

9. Choi JG, Winn AN, Skandari MR, Franco MI, Staab EM, Alexander J, et al. First-Line Therapy for Type 2 Diabetes With Sodium–Glucose Cotransporter-2 Inhibitors and Glucagon-Like Peptide-1 Receptor Agonists. Annals of Internal Medicine [Internet]. 2022 Oct 4 [cited 2023 Feb 9]; Available from: https://www.acpjournals.org/doi/10.7326/M21-2941

10. Drummond MF, Sculpher MJ, Claxton K, Stoddart, Torrance GW. Methods for the economic evaluation of health care programmes 4th edition. 2015.

11. Briggs A, Claxton K, Sculpher and M. Decision Modelling for Health Economic Evaluation. Oxford, New York: Oxford University Press; 2006. 256 p. (Handbooks in Health Economic Evaluation).

12. Hong D. Newer antihyperglycaemic drugs cost effective. PharmacoEconomics & Outcomes News. 2019;824:19–23.

13. Rahman W, Solinsky PJ, Munir KM, Lamos EM. Pharmacoeconomic evaluation of sodium-glucose transporter-2 (SGLT2) inhibitors for the treatment of type 2 diabetes. Expert Opin Pharmacother. 2019 Feb;20(2):151–61.

14. Yoshida Y, Cheng X, Shao H, Fonseca VA, Shi L. A systematic review of cost-effectiveness of sodium-glucose cotransporter inhibitors for type 2 diabetes. Current Diabetes Reports. 2020;20(4):1–19.

15. Bagepally BS, Chaikledkaew U, Gurav YK, Anothaisintawee T, Youngkong S, Chaiyakunapruk N, et al. Glucagon-like peptide 1 agonists for treatment of patients with type 2 diabetes who fail metformin monotherapy: Systematic review and meta-analysis of economic evaluation studies. BMJ Open Diabetes Res Care. 2020 Jul;8(1).

16. Ruan Z, Zou H, Lei Q, Ung COL, Shi H, Hu H. Pharmacoeconomic evaluation of dipeptidyl peptidase-4 inhibitors for the treatment of type 2 diabetes mellitus: A systematic literature review. Expert Review of Pharmacoeconomics & Outcomes Research. 2022;1–20.

17. Zozaya N, Capel M, Simón S, Soto-González A. A systematic review of economic evaluations in non-insulin antidiabetic treatments for patients with type 2 diabetes mellitus. Global & Regional Health Technology Assessment. 2019;2019:2284240319876574.

18. Laursen HVB, Jørgensen EP, Vestergaard P, Ehlers LH. A Systematic Review of Cost-Effectiveness Studies of Newer Non-Insulin Antidiabetic Drugs: Trends in Decision-Analytical Models for Modelling of Type 2 Diabetes Mellitus. PharmacoEconomics [Internet]. 2023 Jul 6 [cited 2023 Aug 8]; Available from: https://link.springer.com/10.1007/s40273-023-01268-5

19. Schmidt M, Schmidt SAJ, Sandegaard JL, Ehrenstein V, Pedersen L, Sørensen HT. The Danish National Patient Registry: A review of content, data quality, and research potential. Clin Epidemiol. 2015;7:449–90.

20. Schmidt M, Schmidt SAJ, Adelborg K, Sundbøll J, Laugesen K, Ehrenstein V, et al. The Danish health care system and epidemiological research: From health care contacts to database records. Clin Epidemiol [Internet]. 2019 Jul 12 [cited 2023 Dec 7];11:563–91. Available from: https://www.ncbi.nlm.nih.gov/pmc/articles/PMC6634267/

21. Kildemoes HW, Sørensen HT, Hallas J. The Danish National Prescription Registry. Scand J Public Health. 2011 Jul;39:38–41.

22. Nargesi AA, Clark C, Aminorroaya A, Chen L, Liu M, Reddy A, et al. Persistence on Novel Cardioprotective Antihyperglycemic Therapies in the United States. The American Journal of Cardiology [Internet]. 2023 Jun 1 [cited 2024 May 20];196:89–98. Available from: https://www.sciencedirect.com/science/article/pii/S0002914923001248

23. Jung H, Tittel SR, Schloot NC, Heitmann E, Otto T, Lebrec J, et al. Clinical characteristics, treatment patterns, and persistence in individuals with type 2 diabetes initiating a glucagon-like peptide-1 receptor agonist: A retrospective analysis of the Diabetes Prospective Follow-Up Registry. DIABETES OBESITY & METABOLISM. 2023 JUL 2023;25(7):1813–22.

24. McGovern A, Hinton W, Calderara S, Munro N, Whyte M, de Lusignan S. A Class Comparison of Medication Persistence in People with Type 2 Diabetes: A Retrospective Observational Study. Diabetes Ther [Internet]. 2018 Feb 1 [cited 2024 May 20];9(1):229–42. Available from: 10.1007/s13300-017-0361-5

25. Kostev K, Gölz S, Scholz B-M, Kaiser M, Pscherer S. Time to Insulin Initiation in Type 2 Diabetes Patients in 2010/2011 and 2016/2017 in Germany. J Diabetes Sci Technol [Internet]. 2019 Nov 1 [cited 2024 May 20];13(6):1129–34. Available from: 10.1177/1932296819835196

26. Poonawalla IB, Bowe AT, Tindal MC, Meah YA, Schwab P. A real-world comparison of cardiovascular, medical and costs outcomes in new users of SGLT2 inhibitors versus GLP-1 agonists. Diabetes Research and Clinical Practice [Internet]. 2021 May [cited 2023 Aug 31];175:108800. Available from: https://linkinghub.elsevier.com/retrieve/pii/S0168822721001595

27. Asche CV, Hippler SE, Eurich DT. Review of Models Used in Economic Analyses of New Oral Treatments for Type 2 Diabetes Mellitus. PharmacoEconomics [Internet]. 2014 Jan [cited 2022 Jul 27];32(1):15–27. Available from: http://link.springer.com/10.1007/s40273-013-0117-7

28. Daly MJ, Elvidge J, Chantler T, Dawoud D. A Review of Economic Models Submitted to NICE’s Technology Appraisal Programme, for Treatments of T1DM & T2DM. Front Pharmacol. 2022;13:887298.

