## Supplemental Files for "Health economic model assumptions of pharmaceutical treatment paths compared with real-world evidence for patients with type 2 diabetes: A nationwide cohort study"

### A1 - Table for CVD codes and variables

*Table of included diagnoses for determining history of cardiovascular disease.*

| Variables | Codes | Notes |
|---|---|---|
| Age | | Derived from days since birth to first use of A10B |
| Sex | | Sex assigned at birth |
| Late | | Whether individual was included in early (2012-2018) or late (2019-2021) cohort |
| Exposure group | | Determined by which of the following drug was first: DPP4 or GLP1 or SGLT2. |
| Duration of diabetes, years | | Time from first NIAD to date of first use of DPP4 or GLP1 or SGLT2 |
| Cardiovascular disease prior to baseline (main cohort) | | Presence of hospitalization with these diagnoses before their index date (first date of NIAD prescription) |
| Cardiovascular disease prior to baseline (nested cohort) | | Presence of hospitalization with these diagnoses before their index date (first date of DPP4/GLP1/SGLT2) |
| Atrial fibrillation | I48 | |
| Hypertension | Hospital codes ICD-10: I10-I15 or use of medicine ATC: C02,C03, C07, C08, C09 | |
| Diabetes with chronic complications | E10.2-E10.8, E11.2-E11.8 | |
| Peripheral vascular disease | I70-I74 or I77 | |
| Prior AMI | I21-I23 | |
| Prior Atherosclerotic cardiovascular disease (ASCVD) | I21, I23, I24, T822A, T823, G45, I20, I25, G45, I672, I678, I679, I691, I693, | |
| | I694, I695, I696, I697, I698, I708, I61, I63, I64, I65, I66, I702, I742, I743, | |
| | I744, I745, I739A, I739C, E105, | |

| Variables | Codes | Notes |
| --- | --- | --- |
| | E115, E145, I700, I739, I748, I749, I709, I740, I741, | |
| Prior PAD | I74.3, I74.5, I70.20, I73.9, E10.5, E11.5,E13.5, E14.5, I70.21, L89, L97, L98.4, M86, R02 | |
| Former TIA | G45 without G45.3 and G45.4 | |

#### A2 - baseline characteristics for main cohort

*Baseline characteristics for the main cohort that was used for treatment pathways*

| | Overall | Early | | Late | Late |
| --- | --- | --- | --- | --- | --- |
| | | Women | Men | Women | Men |
| n | 62,238 | 19,235 | 30,026 | 5,177 | 7,800 |
| Age (mean (SD)) | 57.36 (12.82) | 57.39 (13.62) | 57.30 (12.05) | 57.59 (14.03) | 57.40 (12.83) |
| Age category (%) | | | | | |
| 18-29 | 1,491 (2.4) | 667 (3.5) | 449 (1.5) | 217 (4.2) | 158 (2.0) |
| 30-44 | 8,494 (13.6) | 2,621 (13.6) | 4,104 (13.7) | 646 (12.5) | 1,123 (14.4) |
| 45-59 | 25,983 (41.7) | 7,666 (39.9) | 12,917 (43.0) | 2,104 (40.6) | 3,296 (42.3) |
| 60-74 | 21,141 (34.0) | 6,440 (33.5) | 10,517 (35.0) | 1,641 (31.7) | 2,543 (32.6) |
| 75+ | 5,129 (8.2) | 1,841 (9.6) | 2,039 (6.8) | 569 (11.0) | 680 (8.7) |

#### A3 - Most frequent paths

*The 20 most frequent treatment regimen pathways for the cohort, with the count (n) and percentage (%) of all individuals who experienced each pathway, of the total amount of individuals (N) in each inclusion period. MET = Metformin, DPP4 = dipeptidyl peptidase-4 inhibitor, SGLT2 = sodium-glucose cotransporter 2 inhibitor, GLP1 = glucagon-like peptide 1 receptor agonist, semaglutide-i = injectable semaglutide, semaglutide-o = oral semaglutide, none = above assumption thresholds for continued treatment, so no treatment in a period.*

| Time period | | | | | |
| --- | --- | --- | --- | --- | --- |
| Early (N = 49261) | | | Late (N = 12977) | | |
| Paths | n | % | Paths | n | % |
| MET→SGLT2+MET→MET | 1,925 | 3.91 | MET→GLP1+MET→GLP1 | 1,237 | 9.53 |
| MET→SGLT2+MET | 1,757 | 3.57 | MET→SGLT2+MET | 980 | 7.55 |
| MET→GLP1+MET→GLP1 | 1,679 | 3.41 | MET→SGLT2+MET→MET | 950 | 7.32 |
| MET→METANDDPP4 | 1,473 | 2.99 | MET→SGLT2+MET→SGLT | 912 | 7.03 |

| Time period | | | | | |
|---|---|---|---|---|---|
| | | | 2 | | |
| MET→SGLT2+MET→SGLT2 | 1,376 | 2.79 | MET→GLP1+MET | 615 | 4.74 |
| MET→GLP1+MET | 878 | 1.78 | MET | 563 | 4.34 |
| MET→GLP1+MET→MET | 856 | 1.74 | MET→GLP1+MET→MET | 558 | 4.30 |
| MET→DPP4+MET→MET | 850 | 1.73 | MET→SGLT2 | 512 | 3.95 |
| MET→DPP4+MET→DPP4 | 670 | 1.36 | MET→GLP1 | 469 | 3.61 |
| MET→DPP4 | 635 | 1.29 | MET→METANDDPP4 | 222 | 1.71 |
| MET→DPP4+MET | 572 | 1.16 | MET→DPP4+MET→DPP4 | 207 | 1.60 |
| MET→SU+MET→MET | 559 | 1.13 | MET→None→GLP1 | 206 | 1.59 |
| MET→METANDSGLT2 | 507 | 1.03 | MET→METANDSGLT2 | 182 | 1.40 |
| MET→SGLT2 | 444 | 0.90 | MET→DPP4 | 175 | 1.35 |
| MET→GLP1 | 411 | 0.83 | MET→None→SGLT2 | 174 | 1.34 |
| MET | 404 | 0.82 | MET→DPP4+MET→MET | 140 | 1.08 |
| MET→None→GLP1 | 396 | 0.80 | MET→DPP4+MET | 117 | 0.90 |
| MET→SU+MET | 312 | 0.63 | SGLT2+MET | 106 | 0.82 |
| MET→SU+MET→SU | 304 | 0.62 | SGLT2+MET→MET | 102 | 0.79 |
| MET→None→DPP4 | 292 | 0.59 | GLP1+MET→GLP1 | 83 | 0.64 |

#### A4 - baseline characteristics for nested cohort

*The characteristics of the nested cohort at baseline. Abbreviations: Early/Late = period of inclusion for cohort which was before and after 2019, respectively. DPP4 = dipeptidyl peptidase-4 inhibitors, GLP1 = Glucagon-like peptide-1 receptor agonists, SGLT2 = sodium glucose co-transporter-2, insulin = Insulin, CVD = Cardiovascular disease*

| | | Early | | | | | | | Late | | | | | | |
|---|---|---|---|---|---|---|---|---|---|---|---|---|---|---|---|
| | Overall | Early overall | Female | | | Male | | | Late overall | Female | | | Male | | |
| | | | DPP4 | GLP1 | SGLT2 | DPP4 | GLP1 | SGLT2 | | DPP4 | GLP1 | SGLT2 | DPP4 | GLP1 | SGLT2 |
| n (%) | 55,430 | 24,443 | 5,979 (24 | 1,508 ( 6. | 1,991 ( 8. | 9,694 (39 | 1,776 ( 7. | 34,95 (14 | 30,987 | 2,675 ( 8. | 4,230 (13 | 5,468 (17 | 3,931 (12 | 4,722 (15 | 9,961 (32 |

| | | Early | | | | | | | Late | | | | | | |
|---|---|---|---|---|---|---|---|---|---|---|---|---|---|---|---|
| | | | .5) | 2) | 1) | .7) | 3) | .3) | | 6) | .7) | .6) | .7) | .2) | .1) |
| Age (mean (SD)) | 59.74 (12.89) | 58.88 (12.80) | 61.56 (13.48) | 51.92 (12.94) | 57.84 (12.22) | 59.87 (12.33) | 54.18 (12.06) | 57.54 (11.54) | 60.42 (12.92) | 65.23 (13.38) | 55.38 (14.00) | 61.27 (12.42) | 63.22 (13.10) | 57.47 (12.32) | 61.08 (11.78) |
| Age category (%) | | | | | | | | | | | | | | | |
| 18-29 | 9,73 (1.8) | 426 (1.7) | 79 (1.3) | 109 (7.2) | 43 (2.2) | 79 (0.8) | 64 (3.6) | 52 (1.5) | 547 (1.8) | 26 (1.0) | 260 (6.1) | 80 (1.5) | 24 (0.6) | 75 (1.6) | 82 (0.8) |
| 30-44 | 5,841 (10.5) | 2,847 (11.6) | 564 (9.4) | 298 (19.8) | 250 (12.6) | 987 (10.2) | 322 (18.1) | 426 (12.2) | 2,994 (9.7) | 147 (5.5) | 625 (14.8) | 428 (7.8) | 313 (8.0) | 669 (14.2) | 812 (8.2) |
| 45-59 | 21,223 (38.3) | 9,819 (40.2) | 2,117 (35.4) | 720 (47.7) | 821 (41.2) | 3,840 (39.6) | 807 (45.4) | 1,514 (43.3) | 11,404 (36.8) | 789 (29.5) | 1,725 (40.8) | 1,976 (36.1) | 1,273 (32.4) | 1,952 (41.3) | 3,689 (37.0) |
| 60-74 | 20,734 (37.4) | 8,803 (36.0) | 2,176 (36.4) | 334 (22.1) | 741 (37.2) | 3,713 (38.3) | 531 (29.9) | 1,308 (37.4) | 11931 (38.5) | 1,026 (38.4) | 1,327 (31.4) | 2,238 (40.9) | 1,523 (38.7) | 1,670 (35.4) | 4,147 (41.6) |
| 75+ | 6,659 (12.0) | 2,548 (10.4) | 1,043 (17.4) | 47 (3.1) | 136 (6.8) | 1,075 (11.1) | 52 (2.9) | 195 (5.6) | 4,111 (13.3) | 687 (25.7) | 293 (6.9) | 746 (13.6) | 798 (20.3) | 356 (7.5) | 1,231 (12.4) |
| History of CVD | 4,730 (8.5) | 1,925 (7.9) | 388 (6.5) | 63 (4.2) | 100 (5.0) | 901 (9.3) | 159 (9.0) | 314 (9.0) | 2,805 (9.1) | 193 (7.2) | 145 (3.4) | 411 (7.5) | 396 (10.1) | 354 (7.5) | 1,306 (13.1) |
| Insulin initiation | 1,690 (3.0) | 4,876 (19.9) | 1,346 (22.5) | 335 (22.2) | 265 (13.3) | 2,071 (21.4) | 427 (24.0) | 432 (12.4) | 1,274 (4.1) | 201 (7.5) | 112 (2.6) | 218 (4.0) | 256 (6.5) | 183 (3.9) | 304 (3.1) |
| Diabetes | 2.5 | 1.9 | 1.8 | 1.8 | 2.2 | 1.8 | 1.8 | 2.2 | 3.0 | 3.2 | 3.0 | 3.0 | 3.2 | 2.7 | 3.1 |

| | | | | | | | | | | | | | | | |
|---|---|---|---|---|---|---|---|---|---|---|---|---|---|---|---|
| | | Early | | | | | | | Late | | | | | | |
| duration (mean (SD)) | 7 (2.31) | 2 (1.71) | 1 (1.67) | 7 (1.72) | 4 (1.85) | 3 (1.64) | 1 (1.67) | 4 (1.85) | 8 (2.58) | 7 (2.64) | 2 (2.61) | 9 (2.61) | 4 (2.53) | 7 (2.46) | 2 (2.59) |
| Followup (mean (SD)) | 2.50 (2.12) | 4.17 (2.08) | 4.24 (2.24) | 4.07 (2.18) | 3.77 (1.49) | 4.4 (2.2) | 4.16 (2.21) | 3.72 (1.42) | 1.19 (0.83) | 1.4 (0.86) | 1 (0.76) | 1.17 (0.82) | 1.47 (0.86) | 1.07 (0.78) | 1.18 (0.81) |

#### A5 - CIF times and predictions

Below is the output of observed (CumInc) and predicted cumulative incidence (Pred CumINC). The output is made with the `summary()` function which does not correctly display the amount of events (N events) and the lost-to-follow-up (N lost) counts when the output is in integer intervals.

*Early inclusion. See method section of manuscript for the equation used for prediction. CumInc = cumilative incidence from cumulative incidence function, Pred CumInc = predicted cumulative incidence, SE = standard error, CI = confidence interval. Table is output from R `prodlim` package run through the base R `summary()` function.*

| GROUP | Time (years) | Event | N at risk | N events | N lost | CumInc | SE of CumInc | Lower 95% CI | Upper 95% CI | Pred Cuminc |
|---|---|---|---|---|---|---|---|---|---|---|
| DPP4 | 0 | 1 | 15673 | 0 | 0 | 0.00% | 0.00% | 0.00% | 0.00% | 0.00% |
| DPP4 | 1 | 1 | 14114 | 2 | 2 | 6.79% | 0.20% | 6.39% | 7.19% | 6.79% |
| DPP4 | 2 | 1 | 13086 | 2 | 1 | 11.13% | 0.25% | 10.63% | 11.63% | 11.13% |
| DPP4 | 3 | 1 | 11952 | 1 | 5 | 15.10% | 0.29% | 14.53% | 15.67% | 15.10% |
| DPP4 | 4 | 1 | 8760 | 2 | 7 | 18.77% | 0.33% | 18.13% | 19.41% | 18.77% |
| D | 5 | 1 | 5827 | 0 | 10 | 22. | 0.36% | 21.36% | 22.79% | 22.08% |

| GROUP | Time (years) | Event | N at risk | N events | N lost | CumInc | SE of CumInc | Lower 95% CI | Upper 95% CI | Pred Cuminc |
|---|---|---|---|---|---|---|---|---|---|---|
| PP4 | | | | | | 08% | | | | |
| DPP4 | 6 | 1 | 3547 | 0 | 4 | 25.13% | 0.41% | 24.32% | 25.95% | 25.13% |
| DPP4 | 7 | 1 | 1904 | 0 | 9 | 28.27% | 0.49% | 27.31% | 29.24% | 28.27% |
| DPP4 | 8 | 1 | 877 | 0 | 1 | 30.85% | 0.60% | 29.67% | 32.03% | 32.02% |
| DPP4 | 9 | 1 | 261 | 0 | 2 | 33.12% | 0.75% | 31.65% | 34.59% | 35.22% |
| DPP4 | 10 | 1 | 0 | 0 | 0 | | | | | 38.27% |
| DPP4 | 11 | 1 | 0 | 0 | 0 | | | | | 41.18% |
| DPP4 | 12 | 1 | 0 | 0 | 0 | | | | | 43.95% |
| DPP4 | 13 | 1 | 0 | 0 | 0 | | | | | 46.59% |
| GLP1 | 0 | 1 | 3284 | 0 | 0 | 0.00% | 0.00% | 0.00% | 0.00% | 0.00% |
| GLP1 | 1 | 1 | 2956 | 0 | 0 | 7.03% | 0.45% | 6.15% | 7.91% | 7.03% |
| GLP1 | 2 | 1 | 2712 | 0 | 0 | 12.44% | 0.58% | 11.29% | 13.58% | 12.44% |
| G | 3 | 1 | 2457 | 2 | 3 | 16. | 0.66% | 15.11% | 17.70% | 16.40% |

| GROUP | Time (years) | Event | N at risk | N events | N lost | CumInc | SE of CumInc | Lower 95% CI | Upper 95% CI | Pred Cuminc |
|---|---|---|---|---|---|---|---|---|---|---|
| LP1 | | | | | | 40% | | | | |
| GLP1 | 4 | 1 | 1556 | 1 | 1 | 20.76% | 0.75% | 19.28% | 22.23% | 20.76% |
| GLP1 | 5 | 1 | 1007 | 0 | 2 | 24.19% | 0.85% | 22.52% | 25.85% | 24.19% |
| GLP1 | 6 | 1 | 632 | 0 | 1 | 28.11% | 1.00% | 26.16% | 30.07% | 28.11% |
| GLP1 | 7 | 1 | 365 | 0 | 1 | 30.82% | 1.14% | 28.58% | 33.06% | 30.82% |
| GLP1 | 8 | 1 | 209 | 0 | 0 | 33.83% | 1.37% | 31.16% | 36.51% | 35.60% |
| GLP1 | 9 | 1 | 67 | 0 | 0 | 37.19% | 1.82% | 33.63% | 40.76% | 39.05% |
| GLP1 | 10 | 1 | 0 | 0 | 0 | | | | | 42.31% |
| GLP1 | 11 | 1 | 0 | 0 | 0 | | | | | 45.40% |
| GLP1 | 12 | 1 | 0 | 0 | 0 | | | | | 48.32% |
| GLP1 | 13 | 1 | 0 | 0 | 0 | | | | | 51.09% |
| SGLT2 | 0 | 1 | 5486 | 0 | 0 | 0.00% | 0.00% | 0.00% | 0.00% | 0.00% |

| GROUP | Time (years) | Event | N at risk | N events | N lost | CumInc | SE of CumInc | Lower 95% CI | Upper 95% CI | Pred Cuminc |
|---|---|---|---|---|---|---|---|---|---|---|
| SGLT2 | 1 | 1 | 5181 | 1 | 0 | 3.42% | 0.25% | 2.94% | 3.90% | 3.42% |
| SGLT2 | 2 | 1 | 4889 | 2 | 0 | 6.88% | 0.35% | 6.21% | 7.56% | 6.88% |
| SGLT2 | 3 | 1 | 4388 | 0 | 12 | 9.77% | 0.41% | 8.97% | 10.57% | 9.77% |
| SGLT2 | 4 | 1 | 2105 | 0 | 4 | 12.43% | 0.48% | 11.48% | 13.38% | 12.43% |
| SGLT2 | 5 | 1 | 877 | 0 | 3 | 15.74% | 0.65% | 14.47% | 17.02% | 15.74% |
| SGLT2 | 6 | 1 | 291 | 0 | 1 | 18.98% | 0.94% | 17.13% | 20.83% | 18.05% |
| SGLT2 | 7 | 1 | 118 | 0 | 1 | 21.26% | 1.32% | 18.68% | 23.84% | 20.73% |
| SGLT2 | 8 | 1 | 38 | 0 | 0 | 23.93% | 2.01% | 19.99% | 27.86% | 23.32% |
| SGLT2 | 9 | 1 | 0 | 0 | 0 | | | | | 25.82% |
| S | 10 | 1 | 0 | 0 | 0 | | | | | 28.24% |

| GROUP | Time (years) | Event | N at risk | N events | N lost | CumInc | SE of CumInc | Lower 95% CI | Upper 95% CI | Pred Cuminc |
|---|---|---|---|---|---|---|---|---|---|---|
| GLT2 | | | | | | | | | | |
| SGLT2 | 11 | 1 | 0 | 0 | 0 | | | | | 30.58% |
| SGLT2 | 12 | 1 | 0 | 0 | 0 | | | | | 32.85% |
| SGLT2 | 13 | 1 | 0 | 0 | 0 | | | | | 35.04% |

*Late inclusion. See method section of manuscript for the equation used for prediction. CumInc = cumilative incidence from cumulative incidence function, Pred CumInc = predicted cumulative incidence, SE = standard error, CI = confidence interval. Table is output from R `prodlim` package run through the base R `summary()` function.*

| GROUP | Time (years) | Event | N at risk | N events | N lost | CumInc | SE of CumInc | Lower 95%CI | Upper 95% CI | Pred Cuminc |
|---|---|---|---|---|---|---|---|---|---|---|
| DPP4 | 0 | 1 | 6606 | 0 | 0 | 0.00% | 0.00% | 0.00% | 0.00% | 0.00% |
| DPP4 | 1 | 1 | 4233 | 0 | 10 | 4.93% | 0.29% | 4.37% | 5.50% | 4.93% |
| DPP4 | 2 | 1 | 2075 | 0 | 6 | 8.85% | 0.44% | 7.99% | 9.71% | 8.85% |
| D | 3 | 1 | 0 | 0 | 0 | | | | | 12.98% |

| GROUP | Time (years) | Event | N at risk | N events | N lost | CumInc | SE of CumInc | Lower 95%CI | Upper 95% CI | Pred Cuminc |
|---|---|---|---|---|---|---|---|---|---|---|
| PP4D | 4 | 1 | 0 | 0 | 0 | | | | | 16.92% |
| PP4D | 5 | 1 | 0 | 0 | 0 | | | | | 20.68% |
| PP4D | 6 | 1 | 0 | 0 | 0 | | | | | 24.27% |
| PP4D | 7 | 1 | 0 | 0 | 0 | | | | | 27.70% |
| PP4D | 8 | 1 | 0 | 0 | 0 | | | | | 30.98% |
| PP4D | 9 | 1 | 0 | 0 | 0 | | | | | 34.10% |
| PP4D | 10 | 1 | 0 | 0 | 0 | | | | | 37.08% |
| PP4D | 11 | 1 | 0 | 0 | 0 | | | | | 39.93% |
| PP4D | 12 | 1 | 0 | 0 | 0 | | | | | 42.65% |
| PP4D | 13 | 1 | 0 | 0 | 0 | | | | | 45.25% |
| PP4G | 0 | 1 | 8952 | 0 | 0 | 0.00% | 0.00% | 0.00% | 0.00% | 0.00% |
| LP1G | 1 | 1 | 3916 | 0 | 16 | 3.0 | 0.21% | 2.62% | 3.45% | 3.04% |

| GROUP | Time (years) | Event | N at risk | N events | N lost | CumInc | SE of CumInc | Lower 95%CI | Upper 95% CI | Pred Cuminc |
|---|---|---|---|---|---|---|---|---|---|---|
| LP1 | | | | | | 4% | | | | |
| GLP1 | 2 | 1 | 1327 | 1 | 2 | 6.04% | 0.42% | 5.22% | 6.86% | 6.04% |
| GLP1 | 3 | 1 | 0 | 0 | 0 | | | | | 8.93% |
| GLP1 | 4 | 1 | 0 | 0 | 0 | | | | | 11.72% |
| GLP1 | 5 | 1 | 0 | 0 | 0 | | | | | 14.43% |
| GLP1 | 6 | 1 | 0 | 0 | 0 | | | | | 17.06% |
| GLP1 | 7 | 1 | 0 | 0 | 0 | | | | | 19.60% |
| GLP1 | 8 | 1 | 0 | 0 | 0 | | | | | 22.07% |
| GLP1 | 9 | 1 | 0 | 0 | 0 | | | | | 24.46% |
| GLP1 | 10 | 1 | 0 | 0 | 0 | | | | | 26.78% |
| GLP1 | 11 | 1 | 0 | 0 | 0 | | | | | 29.03% |
| GLP1 | 12 | 1 | 0 | 0 | 0 | | | | | 31.20% |
| G | 13 | 1 | 0 | 0 | 0 | | | | | 33.32% |

| GROUP | Time (years) | Event | N at risk | N events | N lost | CumInc | SE of CumInc | Lower 95%CI | Upper 95% CI | Pred Cuminc |
|---|---|---|---|---|---|---|---|---|---|---|
| LP1 | | | | | | | | | | |
| SGLT2 | 0 | 1 | 15429 | 0 | 0 | 0.00% | 0.00% | 0.00% | 0.00% | 0.00% |
| SGLT2 | 1 | 1 | 7842 | 0 | 24 | 2.84% | 0.15% | 2.54% | 3.14% | 2.84% |
| SGLT2 | 2 | 1 | 3031 | 0 | 9 | 5.29% | 0.26% | 4.78% | 5.81% | 5.29% |
| SGLT2 | 3 | 1 | 0 | 0 | 0 | | | | | 7.84% |
| SGLT2 | 4 | 1 | 0 | 0 | 0 | | | | | 10.31% |
| SGLT2 | 5 | 1 | 0 | 0 | 0 | | | | | 12.72% |
| SGLT2 | 6 | 1 | 0 | 0 | 0 | | | | | 15.06% |
| SGLT2 | 7 | 1 | 0 | 0 | 0 | | | | | 17.34% |
| SGLT | 8 | 1 | 0 | 0 | 0 | | | | | 19.56% |

| GROUP | Time (years) | Event | N at risk | N events | N lost | CumInc | SE of CumInc | Lower 95%CI | Upper 95% CI | Pred Cuminc |
|---|---|---|---|---|---|---|---|---|---|---|
| 2 | | | | | | | | | | |
| SGLT2 | 9 | 1 | 0 | 0 | 0 | | | | | 21.71% |
| SGLT2 | 10 | 1 | 0 | 0 | 0 | | | | | 23.82% |
| SGLT2 | 11 | 1 | 0 | 0 | 0 | | | | | 25.86% |
| SGLT2 | 12 | 1 | 0 | 0 | 0 | | | | | 27.85% |
| SGLT2 | 13 | 1 | 0 | 0 | 0 | | | | | 29.79% |

#### A6 - full cox model

*PY = Person years, IR = Incidence rate, CI = Confidence interval, HR = Hazard ratio, DPP4 = Dipeptidyl peptidase-4 inhibitors, GLP1 = Glucagon-like peptide-1 receptor agonists, SGLT2 = Sodium glucose co-transporter-2, early and late = indicator for inclusion in cohort between 2012-2018 and 2019-2021, respectively. *p<0.1; **p<0.05; ***p<0.01.*

| | (1) | (2) | (3) | (4) |
|---|---|---|---|---|
| GROUP = GLP1 | 1.11 (1.03, 1.19) | 0.64 (0.49, 0.79) | 1.09 (1.01, 1.17) | 0.70 (0.55, 0.85) |
| GROUP = SGLT2 | 0.66 (0.57, 0.74) | 0.59 (0.46, 0.71) | 0.66 (0.58, 0.74) | 0.62 (0.49, 0.74) |
| Male sex | | | 0.93 (0.87, 0.99) | 0.90 (0.79, 1.01) |
| History of CVD | | | 1.46 (1.36, 1.55) | 1.19 (1.01, 1.38) |
| Age | | | 1.00 (1.00, 1.00) | 1.02 (1.01, 1.02) |
| Diabetes Duration | | | 0.97 (0.95, 0.99) | 0.96 (0.94, 0.98) |

| | (1) | (2) | (3) | (4) |
|---|---|---|---|---|
| Time period | Unadjusted early | Unadjusted late | Adjusted early | Adjusted late |
| Observations | 24,443 | 30,987 | 24,443 | 30,987 |
| R2 | 0.01 | 0.002 | 0.01 | 0.004 |
| Max. Possible R2 | 0.98 | 0.55 | 0.98 | 0.55 |
| Log Likelihood | -47,236.59 | -12,397.74 | -47,203.39 | -12,368.92 |
| Wald Test | 121.54 (df = 2) | 74.50 (df = 2) | 191.70 (df = 6) | 133.57 (df = 6) |
| LR Test | 132.89 (df = 2) | 70.81 (df = 2) | 199.31 (df = 6) | 128.46 (df = 6) |
| Score (Logrank) Test | 123.49 (df = 2) | 76.07 (df = 2) | 194.14 (df = 6) | 135.14 (df = 6) |
